# How much reserve capacity is justifiable for hospital pandemic preparedness? A cost-effectiveness analysis for COVID-19 in Germany

**DOI:** 10.1101/2020.07.27.20162743

**Authors:** Afschin Gandjour

**Affiliations:** Frankfurt School of Finance & Management, Frankfurt, Germany

## Abstract

**Introduction:** In preparation for a possible second COVID-19 pandemic wave, expanding intensive care unit (ICU) bed capacity is an important consideration. The purpose of this study was to determine the costs and benefits of this strategy in Germany.

**Methods:** This study compared the provision of additional capacity to no intervention from a societal perspective. A decision model was developed using, e.g., information on age-specific fatality rates, ICU costs and outcomes, and the herd protection threshold. The net monetary benefit (NMB) was calculated based upon the willingness to pay for new medicines for the treatment of cancer, a condition with a similar disease burden in the near term.

**Results:** The marginal cost-effectiveness ratio (MCER) of supplying one additional ICU bed is €24,815 per life year gained and increases with the number of additional beds. The NMB remains positive for utilization rates as low as 1.5% and, assuming full capacity utilization, for multiples of the currently available bed capacity. Expanding the ICU bed capacity by 10,000 beds is projected to result in societal costs of €41 billion and to reduce mortality of ICU candidates by 20% compared with no intervention (assuming full capacity utilization). In a sensitivity analysis, the variables with the highest impact on the MCER were the mortality rates in the ICU and after discharge.

**Conclusions:** In Germany, the provision of additional ICU bed capacity appears to be cost-effective over a large increase in the number of beds. Nevertheless, bed utilization is constrained by labor supply and possibly other input factors.

## Introduction

Following the first severe acute respiratory syndrome coronavirus 2 (SARS-CoV-2) pandemic wave in Germany, the German federal government and the federal states are currently pursuing a strategy of COVID-19 containment. This strategy includes a bundle of measures such as a partial shutdown of businesses, social distancing, tracking, testing, public mask wearing, and quarantine orders. The primary goal of this strategy is to avoid an overproportional increase in the number of new infections by keeping the reproduction number below one. If successful, this strategy will avoid the spread of the virus in the population (up to the point of herd immunity). In addition, this strategy could suppress a second wave of COVID-19 outbreaks or postpone it (‘flatten the curve’) and thus avoid overstretching intensive care unit (ICU) capacity at the time of peak demand. Based on the confirmed COVID-19 cases in Germany as of June 15, 2020, the probability of a COVID-19 patient requiring indication for ICU admission is approximately 8% (Robert Koch Institut 2020).

In general, COVID-19 response measures can be categorized based on the three levels of prevention: primary, secondary, and tertiary prevention (Fletcher 2013). Primary prevention aims to reduce the incidence of COVID-19. Secondary prevention screens asymptomatic and symptomatic patients (with infection) for COVID-19. Tertiary prevention aims to prevent sequelae of COVID-19. While the COVID-19 containment strategy currently pursued by the German government emphasizes primary and secondary prevention, adding ICU bed capacity is an example of tertiary prevention. The German government has pursued the latter strategy until recently, with nearly 9000 beds added as of May 26, 2020 (Bundesamt für Soziale Sicherung 2020), but now intends to redeploy part of the available hospital capacity for treating non-COVID-19 patients (Bundesministerium für Gesundheit 2020). An alternative tertiary prevention strategy that is still under investigation is medical treatment of COVID-19. Currently, there is great hope for future COVID-19 treatments by repurposing drugs that are already approved for other diseases and demonstrate acceptable safety profiles (cf. Kupferschmidt 2020).

The current COVID-19 containment strategy in Germany may turn out to be insufficient in suppressing a possible second SARS-CoV-2 pandemic wave, however. This strategy may also become unsustainable in terms of affordability, psychological burden, or violation of civil rights. Given the renewed risk of an exponential growth of COVID-19 cases, expansion of ICU bed capacity becomes an important consideration again. The purpose of this study was to determine the costs and benefits of this strategy in preparation for a potential second pandemic wave. The objective agrees with a recent recommendation by the German National Academy of Sciences Leopoldina (2020) with regard to the COVID-19 pandemic stating that “[t]he time gained by the shutdown must be used to evaluate the actions taken using empirical data, and the costs and benefits of these actions must be evaluated prior to readjustment”. Results of this study also allow comparing the health benefits and cost-effectiveness of extending ICU bed capacity with those of life extending COVID-19 treatments.

## Methods

### General

I conducted a cost-effectiveness analysis using life-years gained as a measure of health benefits. The analysis was conducted over the remaining lifetime of COVID-19 patients who have an indication for ICU care. By comparing the costs and health benefits of different levels of ICU bed capacity, I calculated the marginal cost-effectiveness ratios (MCERs). In addition, I performed net benefit and return on investment (ROI) calculations.

### Calculation of health benefits

A decision model was constructed using a previously developed and validated model as a basis (Gandjour 2020). The later model determines the loss of life years of a successful shutdown and no intervention, i.e., the situation in which no ICU bed capacity left to treat COVID-19 patients, with each compared with the situation before the pandemic. This paper extends the previous model based on the following conceptual idea: The clinical value of an additional ICU bed is equivalent to the marginal loss of life years in the absence of an additional ICU bed, i.e., when the demand for ICU beds exceeds the available capacity by one ICU bed. Following this principle, I calculated the weighted-average loss of life years when the demand exceeds the available ICU bed capacity by one bed, with weights reflecting the portions of patients admitted to the ICU and refused admission. These weights were multiplied by the average per-capita loss of life years in the German population (compared with non-crisis mortality rates) when all patients with ICU indications were admitted to the ICU and refused admission. The difference in this weighted average compared with the loss of life years with a sufficient ICU bed capacity presents the value of an additional ICU bed. When sequentially adding an *n* number of beds, I applied the same marginal calculation. Given that this calculation relates the addition of beds to the existing national capacity, it was conducted at the population level. For this reason, I multiplied the clinical value of an additional ICU bed by the population size. I conservatively assumed that the benefits of the ICU bed capacity would only last for 12 months, the earliest expected arrival date of a vaccine that protects against COVID-19 (WSJ 2020).

Presuming a harvesting effect in a sensitivity analysis, I assumed for age groups with excess mortality associated with COVID-19 (the difference between observed and pre-pandemic mortality rates) that except for COVID-19, there are no other causes of death in the forthcoming 12 months (Gandjour 2020).

In addition to the above calculation, which yields the clinical value of an ICU bed in terms of life years gained, I also determined its value in terms of reduction of ICU mortality. To this end, I followed the same methodological approach but applied, as weights, mortality of patients admitted to the ICU and refused admission.

### Cost analysis

For the cost analysis, I took a societal viewpoint. To calculate medical costs, I considered the initial ICU stay, rehospitalizations occurring in the first year after discharge from the ICU, hospital copayments, as well as future consumption and unrelated care during added life years. To determine the hospital costs of treating COVID-19 patients, I considered both the operating and infrastructure costs. To calculate the operating costs, I assumed an average patient trajectory. I applied the corresponding diagnosis-related group (DRG) codes plus additional tariffs on top of the DRG payments (“Zusatzentgelt”). Moreover, I considered extra payments by the German government for personal protective equipment and nursing care in treating COVID-19 patients.

To identify the appropriate DRG codes for COVID-19 cases admitted to the ICU, I followed the guidance of the German Interdisciplinary Association for Intensive Care and Emergency Medicine (DIVI 2020). Specifically, I applied DRG codes that reflect the average LOS with and without mechanical ventilation. I purposely used conservative cost estimates, i.e., I selected higher-cost DRGs in the presence of several coding options, thus biasing against the value of an additional ICU bed. To arrive at the final cost estimate for treating a COVID-19 patient in the ICU, the costs of patients with and without mechanical ventilation were weighted by their respective shares.

To arrive at the costs of infrastructure, I accounted for the opportunity costs of capital. To calculate the latter, I considered the weighted average cost of capital (WACC). Strictly speaking, the WACC only applies to private hospitals, which account for 36% of all German hospitals, based on 2016 data (BDPK 2020). However, during the coronavirus crisis government funds cover a portion of the capital costs resulting from the expansion of ICU capacity, i.e., €50,000 per additional ICU bed (Bundesregierung 2020). Hence, the WACC needs to be adjusted for this portion (cf. Zapp 2010). In contrast, when public hospitals expand their capacity, they receive interest-free loans from the federal states without any obligation to pay them back. Nevertheless, only half of the infrastructure investments are currently covered by the federal states (GKV-Spitzenverband 2018). The overall opportunity cost of capital was thus calculated as a weighted average of the WACC and a zero cost of capital, with the weights representing shares of private and public funding, respectively.

To determine the future unrelated medical costs incurred during added life years, I determined the cumulative probability of an individual at age *i* of surviving until age *j* (i.e., the product of age-specific survival probabilities up to age *j*) using the life table embedded in the previously published decision model (Gandjour 2020). I multiplied the cumulative probability of surviving until age *j* by health expenditures at age *j*, took the sum over all ages *j*, and thus obtained the remaining health expenditures for an individual at age *i*. By comparing the remaining health expenditures for different levels of ICU bed capacity, I calculated the life extension costs. To account for the age distribution of the population, I weighted the age-specific life-extension costs by age-specific population sizes. I performed all calculations for men and women separately and then aggregated the results.

Moreover, a societal perspective requires considering expenses for primary needs such as food, shelter, and clothing as their satisfaction contributes to life extension (Gandjour 2006). That is, as the denominator of the MCER captures the benefits of the resources used to satisfy primary needs, the costs of these resources also need to be included for consistency reasons (Gandjour 2006). To determine these types of consumption costs during added life years, I used the same calculation as for health expenditures outlined in the above paragraph.

### Net benefit and ROI calculation

The monetary value of an additional life year was borrowed from new, innovative oncological drugs as cancer reflects a condition with a similar morbidity and mortality burden in the general population in the short term as COVID-19 (Gandjour 2020). To calculate the net monetary benefit (NMB) of an additional bed, I subtracted the cost of an additional bed from the monetary value created. By dividing the monetary value of an additional bed by its additional cost, I also determined the ROI.

### Discounting

In the base-case analysis, I did not discount costs or health benefits, as the reported survival benefits from cancer treatment (Storm 2017), which were used to determine the economic value of a life year, were undiscounted as well. In a sensitivity analysis, I discounted both costs and effects.

### Sensitivity Analysis

Using one-way deterministic analyses, I assessed the parameter uncertainty by varying the input parameters that are subject to variation one at a time. In addition, I conducted threshold sensitivity analyses that determined the break-even points for additional ICU bed capacity, government subsidies for ICU bed provision, and ICU bed utilization.

## Data

The model input data are listed in Table 1. For COVID-19 patients receiving mechanical ventilation, LOS in the ICU has been estimated to be between 11 and 20 days (KSTA 2020, Stang 2020, SWR 2020). Among the DRGs that are applicable in this range, I made conservative choices. Of note, in the German DRG system, age-specific DRG codes are usually limited to children and thus played no role in assigning DRG codes. Specifically, for ICU patients receiving mechanical ventilation, I chose the DRG code E40A (InEK 2020), which is the only DRG code with a specific reference to ARDS. It has a case-mix index of 3.406 and allows for an additional payment of €18.21 (code ZE162). For ICU patients who do not receive mechanical ventilation, I applied the DRG code E77B, which entails a slightly shorter LOS (15.1 versus 17.1 days) with a case-mix index of 2.090 and allows for an additional payment of €34.48 (code ZE163). Each case-mix index was multiplied by the national base price (GKV-Spitzenverband 2020). Of note, the DRG codes I applied cover all hospital expenditures including nursing costs.

**Table 1.** Input data used in the base case and the sensitivity analysis.

| Input | Mean (range) | Reference |
| --- | --- | --- |
| *Epidemiological and clinical data* |  |  |
| Probability of death by age and gender in Germany | see reference | Statistisches Bundesamt 2019 |
| Population size by age | see reference | Statistisches Bundesamt 2020 |
| CFR in Germany |  | Robert Koch Institut 2020 |
| Total population | 0.047 (0.0036 – 0.047) |  |
| 0-9 years | 0.0002 |  |
| 10-19 years | 0.0002 |  |
| 20-49 years | 0.0012 |  |
| 50-69 years | 0.0197 |  |
| 70-89 years | 0.1987 |  |
| 90+ years | 0.3111 |  |
| Probability of ICU indication | 0.076 (0.04 – 0.08) | Robert Koch Institut 2020 |
| CFR in the ICU | 0.30 (0.21 – 0.52) | Robert Koch Institut 2020 |
| CFR for ICU non-admission | 0.25 (0.21 – 0.52) | Robert Koch Institut 2020 |
| CFR one year post ICU discharge | 0.1 (0.1 – 0.2) | Abers 2014 |
| Herd protection threshold | 0.70 (0.60 – 0.70) | Kwok 2020 |
| ICU beds available for COVID-19 | 12,279 | Robert Koch Institut 2020 |
| Weighted average cost of capital | 0.06 | PwC 2020 |
| Proportion of ICU patients with mechanical ventilation | 0.61 | Robert Koch Institut 2020 |
| ICU length of stay | 14 (11 – 20) | KSTA 2020, Stang 2020, SWR 2020 |
| *Cost data* |  |  |
| Healthcare expenditure by age | see reference | Statistisches Bundesamt 2015 |
| Consumption costs per year, primary needs (€) |  | Statistisches Bundesamt 2019 |
| Adult | 11,580 |  |
| Child | 3984 |  |
| ICU bed, infrastructural cost (€) | 85,000 (85,000 – 100,000) |  |
| ICU costs per admission (with mechanical ventilation) (€) | 12,551 | GKV-Spitzenverband 2020, InEK 2020 |
| ICU costs per admission (without mechanical ventilation) (€) | 7725 | GKV-Spitzenverband 2020, InEK 2020 |
| Hospital copayment per day (€) | 10 | Bundesministerium für Gesundheit 2018 |
| Extra payment for nursing care per day (€) | 147 | Bundesregierung 2020 |
| Personal protective equipment per patient (€) | 50 | Bundesregierung 2020 |

ICU = intensive care unit, CFR = case fatality rate

In terms of the infrastructure costs of ICU beds, the estimates range between €85,000 and €100.000 (DIVI 2020, DKG 2020, ZEIT 2020). In the base case, I applied an estimate provided by the German Hospital Federation, which was €85,000 (DKG 2020). This estimate includes the costs of supplies (e.g., protective clothing) and equipment (e.g., ventilators) associated with ICU bed provision (DIVI 2020).

To estimate the costs of rehospitalizations occurring in the first year after discharge from the ICU, I used the results of a published cohort study of 396 ICU survivors with acute respiratory distress syndrome. The study was conducted between September 2014 and April 2016 in 61 German hospitals (Brandstetter 2019). The study reported a median number of rehospitalizations of 2 (interquartile range 1-3). The LOS was 16 days on average (interquartile range 10-25). Of note, rehospitalizations included stays in rehabilitation facilities as well as admissions for medical conditions unrelated to acute respiratory distress syndrome. The data did not differentiate between different types of rehospitalizations or admissions to ICUs and normal wards. Given the latter, I applied the costs of the initial ICU stay, thus conservatively assuming that all rehospitalized patients would be admitted to the ICU.

As outlined in the Methods section, for ICU survivors I determined both future (unrelated) medical and consumption costs during added life years. To account for the former, I used the healthcare expenditures in the general population, which were available for four age categories (Statistisches Bundesamt 2015) and explicitly referred to national (societal) costs and not to social health insurance costs. The data on the private consumption costs were from 2017 (Statistisches Bundesamt 2019). The categories of the private consumption costs were available for adults and children but not according to age. To narrow down the consumption costs to those for primary needs, I considered the per capita private consumption costs on food and nonalcoholic beverages, clothing and shoes, housing (including maintenance), energy, and health.

The capital costs were based on the whole healthcare and pharmaceutical industry (PwC 2020). All costs were inflated to 2020 euros based on the general German Consumer Price Index.

The willingness to pay for an additional life year (€101,493 per life year gained) was obtained by dividing incremental costs of new, innovative cancer drugs (€39,751) by the incremental survival benefit (0.39 life years) (Gandjour 2020).

Given the lack of official guidance on the discount rates for the costs and health benefits from a societal perspective in Germany, I applied a 3% discount rate for the costs based on the social rate of time preference derived from the Ramsey equation (1928). For health benefits, I applied a 1% lower discount rate, reflecting the expected growth rate of the consumption value of health in Germany (cf. John 2019).

## Results

Expanding the existing ICU bed capacity by exactly one bed yields a MCER of €24,815 per life year gained and an ROI of 3.9 in the base case. The cost-effectiveness and ROI diminish with additional increases in ICU bed capacity (Figure 1). This trend holds because when the demand for ICU beds exceeds the available capacity, the marginal loss of life years in the absence of an additional bed diminishes. The reason is that an increase in excess demand leaves a growing share of the patient population without ICU admission and hence, the nonprovision of ICU bed capacity matters relatively less. Hence, each additional ICU bed prevents relatively less excess demand. For a given demand level, adding one bed to 1000 beds at the baseline is thus more valuable than adding one bed to 10,000 beds.

**Figure 1.**
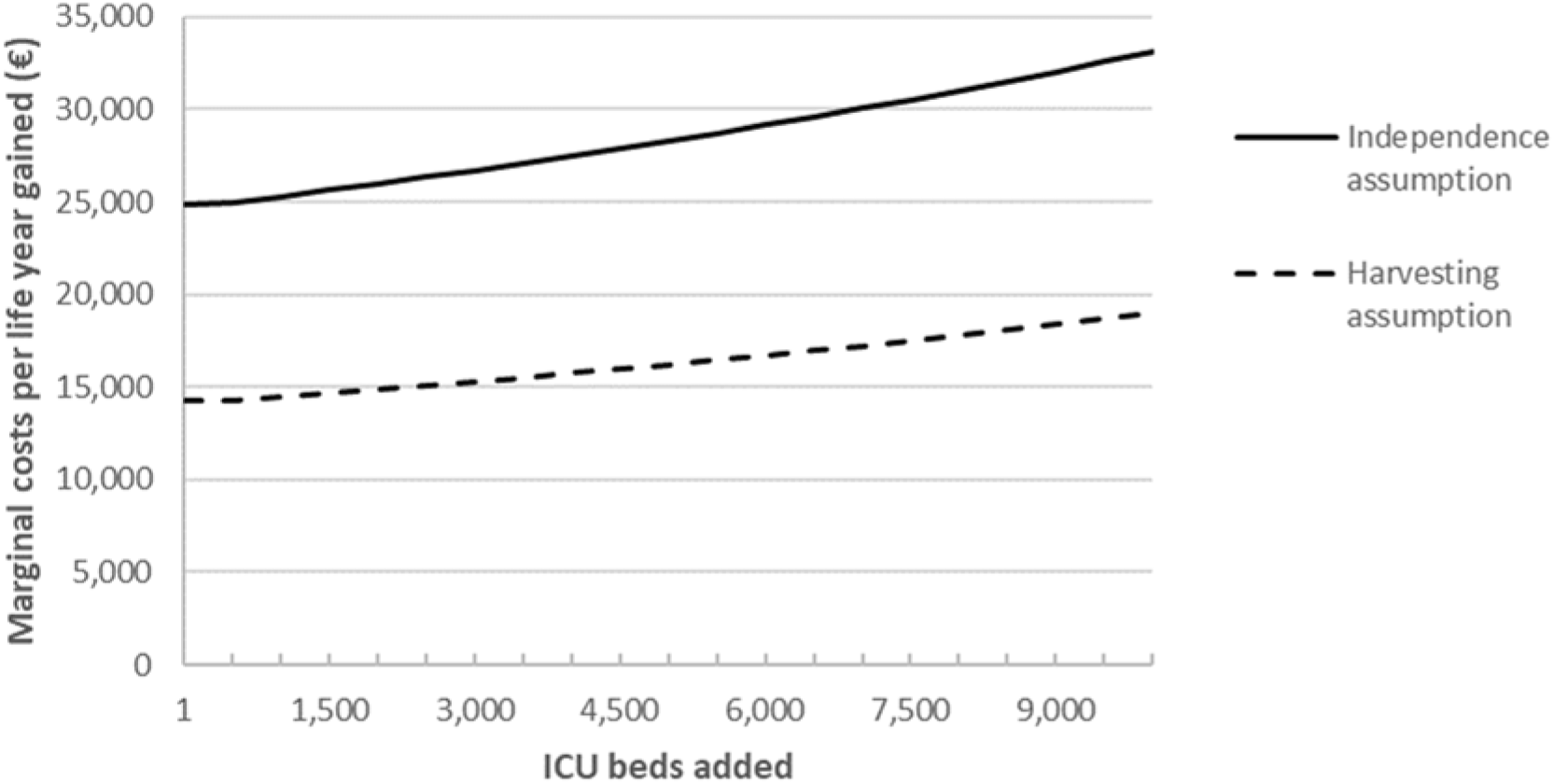
Marginal costs per life year gained by adding intensive care unit (ICU) bed capacity.

Based on the harvesting assumption, the cost-effectiveness of supplying an additional ICU bed improves because COVID-19 patients who are saved from death in the presence of an additional bed are assumed to represent a healthier subgroup of ICU patients than those who unavoidably die.

As shown in the sensitivity analysis (see Figure 2), the variables with the greatest impact on NMB were the mortality rates in the ICU and after discharge. Ceteris paribus, higher mortality rates reduce the NMB of an additional ICU bed.

**Figure 2.**
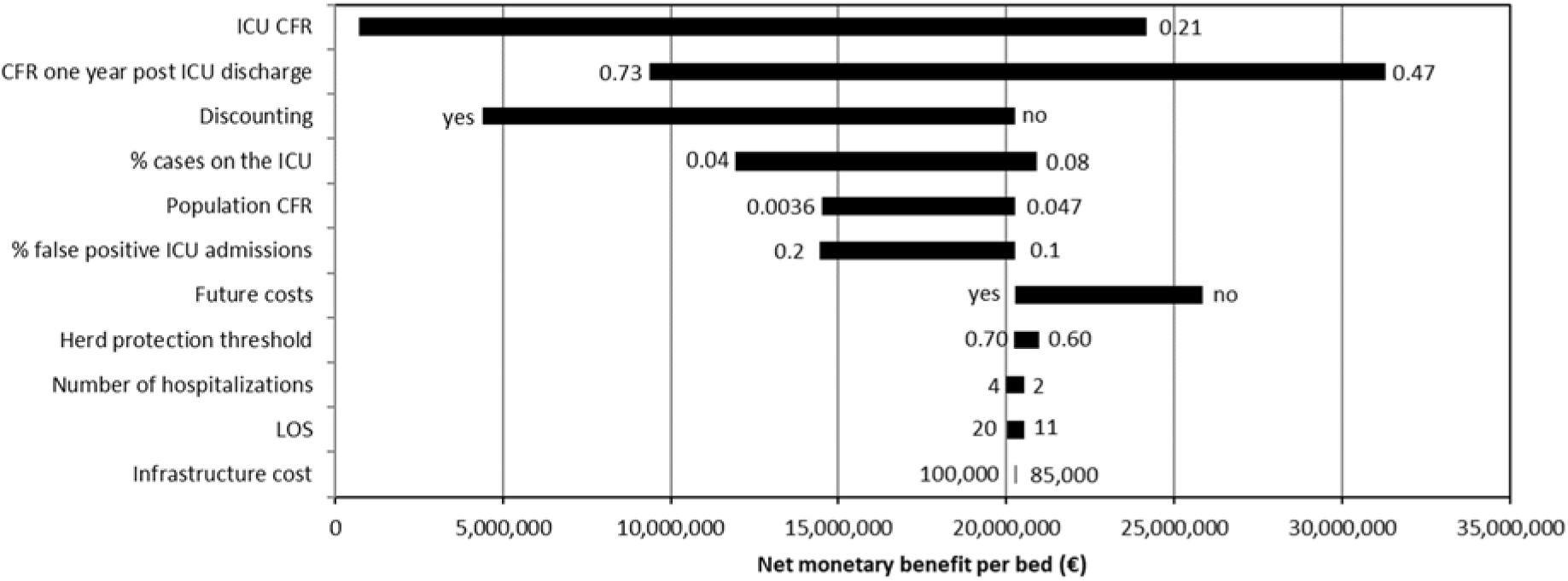
Tornado diagram demonstrating the results of the one-way sensitivity analysis. The variables are ordered by impact on the net monetary benefit of the provision of additional ICU bed capacity versus no intervention. The numbers indicate the upper and lower bounds. ICU = intensive care unit, CFR = case fatality rate, LOS = length of stay

Expanding ICU bed capacity by another 10,000 beds or 81% of the currently available capacity is projected to increase societal costs by €40.6 billion. The resulting decrease in the mortality of ICU candidates is 20% compared with no intervention (Figure 3). While the ROI diminishes with the expansion of capacity, it remains above 3.0 for the ten thousandth bed added. Even a bed utilization of only 1.5% still allows for a positive ROI due to the low share of infrastructure costs.

**Figure 3.**
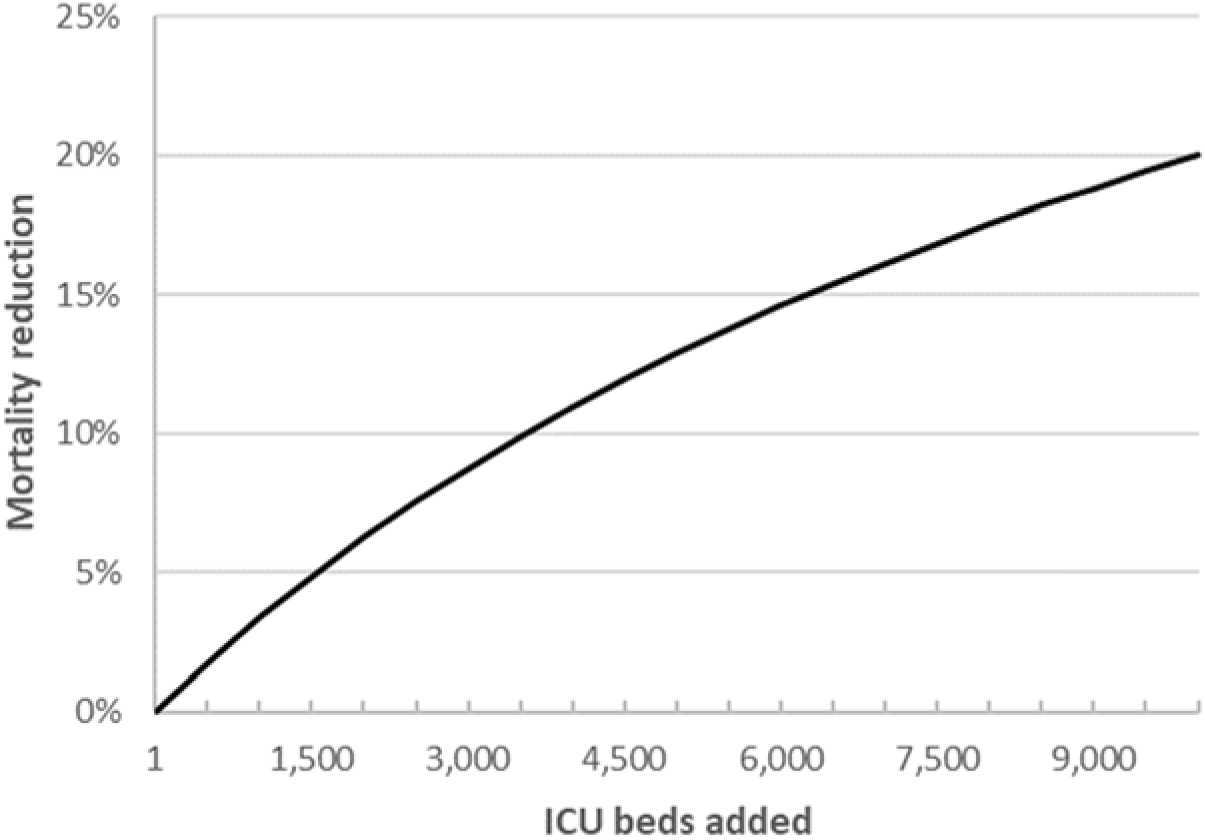
Mortality reduction of intensive care unit (ICU) candidates by increasing ICU bed capacity compared with no increase.

Threshold sensitivity analysis shows that negative returns do not appear even with a 11-fold increase in ICU bed capacity. Similarly, a government subsidy of €5 million per ICU bed still yields a positive NMB for the ten thousandth bed added.

## Discussion

In preparation for a possible second SARS-CoV-2 pandemic wave, the German government has adopted a COVID-19 containment strategy. The current strategy may turn out to be insufficient in preventing overstretching of ICU capacity at the time of peak demand, however. Therefore, the expansion of ICU bed capacity becomes a relevant consideration. As shown in this analysis, increasing ICU bed capacity provides a high ROI over a large increment of beds.

Extending the existing ICU bed capacity seems acceptable based on the MCER but also from a budgetary perspective. That is, extending capacity by more than 81% is forecasted to result in a one-time increase in healthcare expenditure (Statistisches Bundesamt 2020a) of 10%, which amounts to 1.2% of the gross domestic product (GDP) in Germany (Statistisches Bundesamt 2020b). In light of these findings and in particular the threshold sensitivity analysis, the current government subsidy for additional ICU bed provision, which is €50,000 per bed, seems inadequate.

Whether the ROI of ICU bed expansion is larger than that of a shutdown of businesses still needs to be investigated. As a complicating factor, calculating the latter needs to account for the potential underfunding of the health care system as a result of the comparatively larger reduction in the GDP. In case the ROI of ICU bed expansion turns out to be higher, a switch from the current strategy of primary and secondary prevention of COVID-19 to a strategy focusing on secondary and tertiary prevention may still not be warranted. The reason is that an isolated strategy of ICU bed expansion without decrease in infection rates inevitably leads to deaths, sickness, and quarantines of the working population and hence a decrease in GDP secondary to productivity loss.

As a word of caution, cost-effectiveness of ICU bed expansion is based on the assumption of a positive probability of utilizing the additional ICU bed capacity. If, however, the additional capacity remains entirely unused, the value of investment becomes negative due to the presence of fixed costs. It is reassuring, however, that even a vacancy rate of 98% still allows for a positive return due to the low share of infrastructure costs. This is equivalent to a 2% probability of having full utilization. How does this finding fit into the virus containment strategy currently pursued by the German government, which aims to keep the reproduction number below one until a vaccine is available? A strategy of supplying additional ICU beds becomes cost-effective once there is a 2% probability that the virus containment strategy is not successful or is abandoned because it is too expensive or burdensome for society or because a vaccine is not available in time globally. Hence, an economic justification for a bed expansion strategy requires a positive probability of viral spread in the population, thus leading to herd immunity by natural infection, regardless of whether this is actively sought by the government. Nevertheless, treating additional COVID-19 patients in the ICU may require cancelling or postponing unrelated treatments. Considering the latter thus increases the minimum acceptable bed utilization rate. Therefore, a strategy of supplying additional ICU beds still requires careful planning.

Of note, there are different ways of providing the additional ICU capacity. These approaches not only include the construction of new buildings but also freeing up existing capacity, e.g., deferring elective procedures, moving non-COVID-19 patients to alternative sites, and using step down care more aggressively. In addition, ICU units and beds may be converted from existing capacity, such as operating, recovery, procedure and treatment rooms; ambulatory surgery centers; unstaffed floors; physical therapy space; outpatient facilities; and nonhealthcare facilities (Government of Alberta 2020, Singhal 2020). In the short term, freeing up existing capacity may, in fact, be the only feasible approach. To meet a potential increase in future demand for ICU beds, the construction of new buildings and the conversion of existing capacity may be unavoidable, however.

Preparing for an increased demand for ICU care also requires recruiting additional personnel (e.g., ICU nurses) as well as purchasing additional materials, supplies (e.g., protective clothing), and equipment (e.g., ventilators). Strategies to address a shortage of labor include accelerated training for ICU nurses; contacting former nurses with ICU experience and other recently retired staff; and redeploying anesthesiologists, other physicians, other nurses, respiratory therapists, other allied health professionals and other staff with appropriate skills to work in a critical care environment (Government of Alberta 2020). Even before the Sars-CoV-2 pandemic the health care environment in Germany has been characterized by labor shortages, particularly ICU nurses (DGIIN 2019). Hence, the above strategies may not be sufficient and increases in the availability of labor may only be feasible through long-term investment in training and education. In 2018, the German government has taken steps to improve short-term and long-term nurse staffing (BMG 2018).

Recently, dexamethasone was shown to lower the 28-day mortality among hospitalized COVID-19 patients receiving respiratory support (Recovery Collaborative Group 2020). Expanding ICU bed capacity and life-prolonging treatments of COVID-19 become complementary interventions if life-prolonging treatments have a label for ICU patients. In that case, ICU bed expansion becomes an enabling strategy for life-prolonging treatments and creates option value (cf. Garrison 2017). Both interventions applied together thus form a combination therapy. This is also confirmed in the sensitivity analysis of this paper, implying an improved cost-effectiveness of ICU bed expansion when accounting for an ICU mortality reduction of life-prolonging treatments. Of note, the absolute mortality reduction demonstrated by dexamethasone (12%) is already achieved by an ICU capacity expansion of 5000 beds (assuming full capacity utilization), thus emphasizing the clinical significance of ICU bed expansion.

If expanding ICU bed capacity and providing life-prolonging treatments of COVID-19 are complementary approaches but ICU bed capacity is a limiting factor, life-prolonging treatments may not be applicable unless they are prescribed on-label or off-label before ICU admission. In that case, life-prolonging treatments can be regarded as a substitute for expanding ICU bed capacity. Similarly, if life-prolonging treatments have a label for hospitalized patients without mechanical ventilation and are able to reduce ICU admissions, they become a substitute for expanding ICU bed capacity.

The limitations of this study need to be acknowledged. First, there are reasons why the base-case analysis overestimates the NMB, i.e., underestimates the MCER. Some of these reasons were already described in the sensitivity analysis and include high mortality in the ICU and post discharge. Furthermore, this study did not include direct nonmedical costs, such as time and transportation costs, which are mandated by the societal perspective adopted in this paper.

On the other hand, there are reasons to believe that the base case underestimates the NMB, i.e., overestimates the MCER. First, by including future medical costs along with the costs of hospitalizations in the first year after ICU discharge, some double counting of hospitalization costs may result. Furthermore, the DRG rates may not reflect true hospital costs and may yield a positive profit margin for ventilated patients (Welt 2020). Moreover, the productivity gains resulting from a reduction in mortality were not included due to the age distribution of averted deaths (the median age is 82 years) and the difficulty of disentangling deaths in relevant age groups (e.g., in the age group 50-69 years). Some of the biases mentioned in this and the previous paragraph may cancel each other out, however.

In terms of the transferability of the results and conclusions of this study to other countries the usual caveats apply. The reasons for caution include between-country differences in clinical and epidemiological data, costs, and the willingness to pay for health benefits.

For data collection in the forthcoming months of the crisis, policymakers should pay particular attention to mortality data, as the MCERs and ROIs forecasted in this study were shown to be particularly sensitive to these data.

## Data Availability

All data generated or analysed during this study are included in this published article.

## Acknowledgements

I would like to thank Rolf Bartkowski and Rainer Sibbel for valuable comments on the hospital cost calculation. Any remaining errors are, of course, my responsibility.

